# Infection-induced immunity is associated with protection against SARS-CoV-2 infection, but not decreased infectivity during household transmission

**DOI:** 10.1101/2022.10.10.22280915

**Authors:** Aaron M Frutos, Guillermina Kuan, Roger Lopez, Sergio Ojeda, Abigail Shotwell, Nery Sanchez, Saira Saborio, Miguel Plazaola, Carlos Barilla, Eben Kenah, Angel Balmaseda, Aubree Gordon

## Abstract

**Background:** Understanding the impact of infection-induced immunity on SARS-CoV-2 transmission will provide insight into the transition of SARS-CoV-2 to endemicity. Here we estimate the effects of prior infection induced immunity and children on SARS-CoV-2 transmission in households.

**Methods:** We conducted a household cohort study between March 2020-June 2022 in Managua, Nicaragua where when one household member tests positive for SARS-CoV-2, household members are closely monitored for SARS-CoV-2 infection. Using a pairwise survival model, we estimate the association of infection period, age, symptoms, and infection-induced immunity with secondary attack risk.

**Results:** Overall transmission occurred in 72.4% of households, 42% of household contacts were infected and the secondary attack risk was 13.0% (95% CI: 11.7, 14.6). Prior immunity did not impact the probability of transmitting SARS-CoV-2. However, participants with pre-existing infection-induced immunity were half as likely to be infected compared to naïve individuals (RR 0.53, 95% CI: 0.39, 0.72), but this reduction was not observed in children. Likewise, symptomatic infected individuals were more likely to transmit (RR 24.4, 95% CI: 7.8, 76.1); however, symptom presentation was not associated with infectivity of young children. Young children were less likely to transmit SARS-CoV-2 than adults. During the omicron era, infection-induced immunity remained protective against infection.

**Conclusions:** Infection-induced immunity is associated with protection against infection for adults and adolescents. While young children are less infectious, prior infection and asymptomatic presentation did not reduce their infectivity as was seen in adults. As SARS-CoV-2 transitions to endemicity, children may become more important in transmission dynamics.

**Article summary:** Infection-induced immunity protects against SARS-CoV-2 infection for adolescents and adults; however, there was no protection in children. Prior immunity in an infected individual did not impact the probability they will spread SARS-CoV-2 in a household setting.

## Introduction

As SARS-CoV-2 transitions from a pandemic phase to endemicity, more individuals will have infection-induced immunity and children will increasingly represent the greatest proportion of primary infections. [1] Thus, understanding the impact of infection-induced immunity on transmission and contribution of children to SARS-CoV-2 transmission is essential to understanding how this transition will occur.

Prior transmission studies show that vaccination reduces the likelihood of transmission, [2, 3] and infection-induced immunity is associated with shorter shedding duration and lower viral load;[4] however, the effect of infection-induced immunity on SARS-CoV-2 transmission has not been well established.[5] Given the high infectivity of SARS-CoV-2 and its emerging variants, many children have already been infected worldwide.[6-9]. Further, as of June 2022, SARS-CoV-2 vaccine availability and uptake has been limited for children globally.[10]

Questions persist about the contribution of children to SARS-CoV-2 transmission. Evidence on the contribution of children to transmission generally shows that children have a lower risk of SARS-CoV-2 transmission when infected compared to adults [11-13] while other work, particularly after the emergence of SARS-CoV-2 variants, finds that children have similar or increased risk of transmission.[14, 15]

We present results from an ongoing, community-based, household transmission study located in Managua, Nicaragua from June 2020-June 2022. We evaluate the effect of prior infection-induced immunity on transmission as well as the contribution of children to SARS-CoV-2 household transmission.

## Methods

This study was approved by institutional review boards at the Nicaraguan Ministry of Health and the University of Michigan. Adults and parents/guardians of children provided written informed consent and children six years or older provided verbal assent.

Participants included in this analysis are members of the ongoing Household Influenza Cohort Study (HICS) which began in 2017. HICS is a community-based prospective household cohort study located in District II of Managua, Nicaragua. In June 2020, the study was expanded to include a transmission sub-study of SARS-CoV-2. Participants attend the Health Center Sócrates Flores Vivas at the first signs of a fever or respiratory illness. A respiratory sample is collected and tested for influenza and SARS-CoV-2 via reverse-transcription polymerase chain reaction (PCR).

When a participant tests positive for SARS-CoV-2, household members are invited to participate in the SARS-CoV-2 transmission sub-study. A separate consent was collected for the sub-study. Study staff visit the home up to six times to collect respiratory samples (days 0, 3, 7, 14, 21, and 30) and conduct a final follow-up visit 45-60 days later. Daily symptom data is collected by staff during each visit. [16]

Blood samples were collected twice per year and risk factor surveys were collected annually. All blood samples collected from 2020-2021 were tested for SARS-CoV-2 IgG antibody to the spike receptor binding domain via an enzyme-linked immunosorbent assay (ELISA) following a protocol adapted from Mount Sinai.[17]

Prior SARS-CoV-2 infection-induced immunity included both PCR and serologically confirmed infections. We categorize SARS-CoV-2 infections into three periods: March 2020-February 2021 (pre-variant era), March 2021-December 2021 (pre-omicron variants, predominantly gamma and delta), and January 2022-June 2022 (omicron variant).

SARS-CoV-2 vaccinations in the cohort began in 2021. Most vaccinated participants received their first vaccine beginning in September of 2021. A variety of vaccines have been used, with AstraZeneca, Abdala, and the Soberana 02 being the three most common vaccines administered. Participants are considered fully vaccinated 14 days after the final dose. We compared age at enrollment, sex, SARS CoV-2 vaccination, and presence of SARS-CoV-2 antibodies before January 1, 2022, between participants who did and did not participate in intensive monitoring using a chi-square and Fisher-exact tests. Using these tests, we also compared infection period, sex, age, bedroom- and bed-sharing, prior infections, vaccination, and index case symptoms between households that did and did not have transmission (an observed SAR-CoV-2 infection among household members) and (except for symptoms) between PCR- and PCR+ household contacts.

To estimate the household secondary attack risk (SAR) and rate ratios (RR), we used pairwise survival models which estimate failure time based on contact intervals between infectious and susceptible contacts. These models can account simultaneously for within-household transmission and the risk of infection from outside the household. The SAR from these models can be interpreted as the probability of transmission from one infected household member to one susceptible. [18, 19]

We assumed an incubation period of six days, a latency period of three, and a 10-day duration of infectiousness; [20-22] therefore, participants were considered infectious three days before to seven days following infection. Participants were considered symptomatic during their infectious period if they reported symptoms within seven days following the infection date.

SAS version 9.4 (SAS Institute Inc.) and R version 4.1.1 with the transtat package were used to conduct the analysis.[18, 23] The models included infection period, number of household members, age, sex, presence of symptoms, cough, rhinorrhea, prior SARS-CoV-2 infection, SARS-CoV-2 vaccination, and bed- and bedroom-sharing. We also include an interaction term for age with presence of symptoms, cough, rhinorrhea, and prior SARS-CoV-2 infection. To evaluate if the household SARs were different when considering only households infected with the omicron variant, we reran the univariate models for household activation for 2020/2021 and 2022. For sensitivity analyses, we adjusted the assumed incubation, latency, and infectious periods. We also reran the univariate models including only households where all household members consented to participate in the household activation and serial swabbing.

## Results

From March 2020-June 2022, there were 2,398 active participants in the cohort with 84 new/re-enrollees, 251 withdrawn, and 23 deaths (Supplemental Figure 1). Within the SARS-CoV-2 transmission sub-study, a total of 209 households (48% of all cohort houses) were activated (some multiple times) with 297 total activations and 1,189 household contacts that consented to intensive monitoring and 258 that declined participation or were not present. Participants in activated households that did not participate in intensive monitoring were generally working-age adults and male. They also had lower cohort participation, were more likely to have missed cohort blood collections since the start of the pandemic and were less likely to have reported vaccination or have documented SARS-CoV-2 antibodies (Table 1, Supplemental Figure 2). In addition to the 297 primary cases, 494 household contacts (42%) were infected.

**Table 1.**
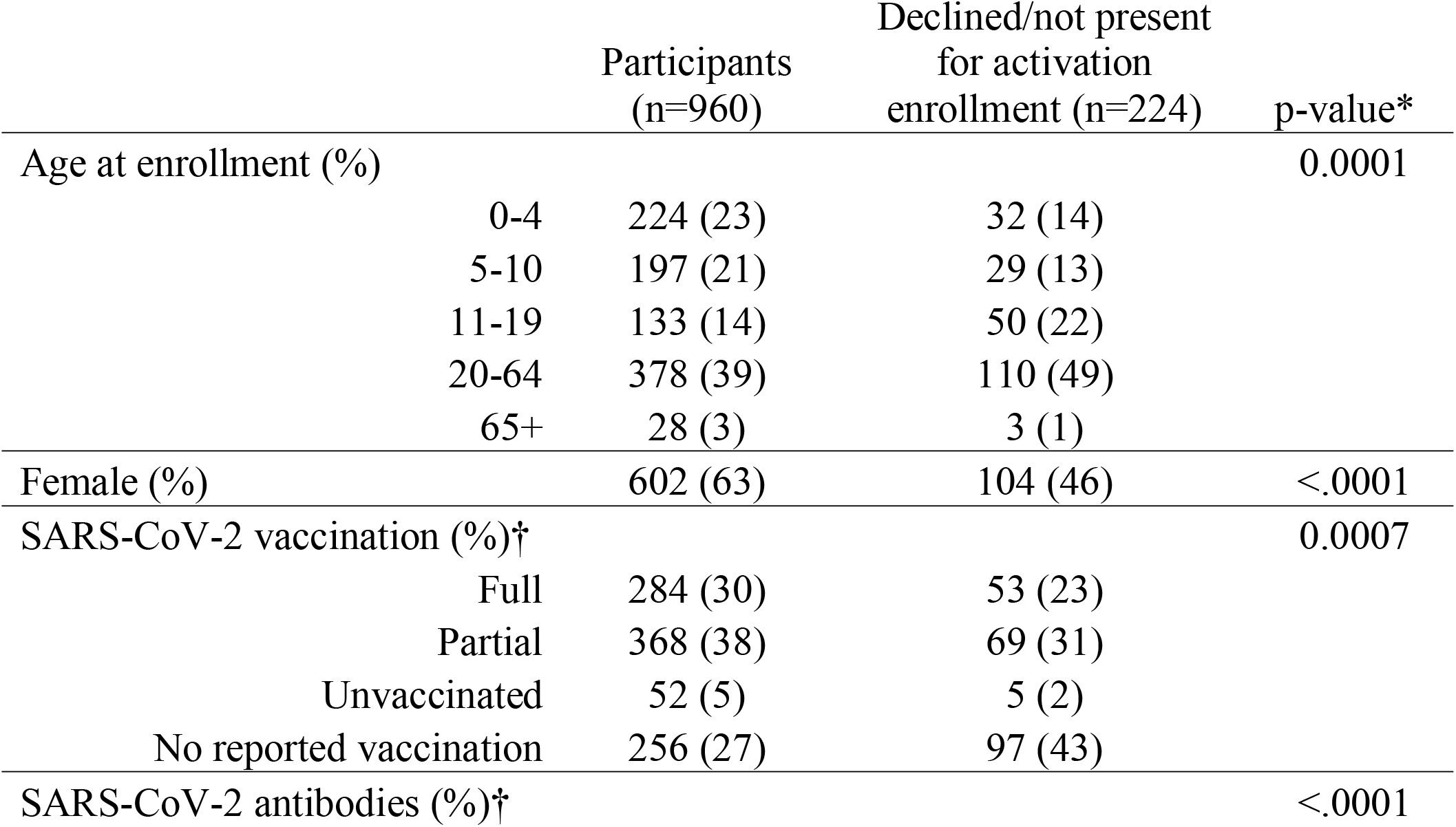
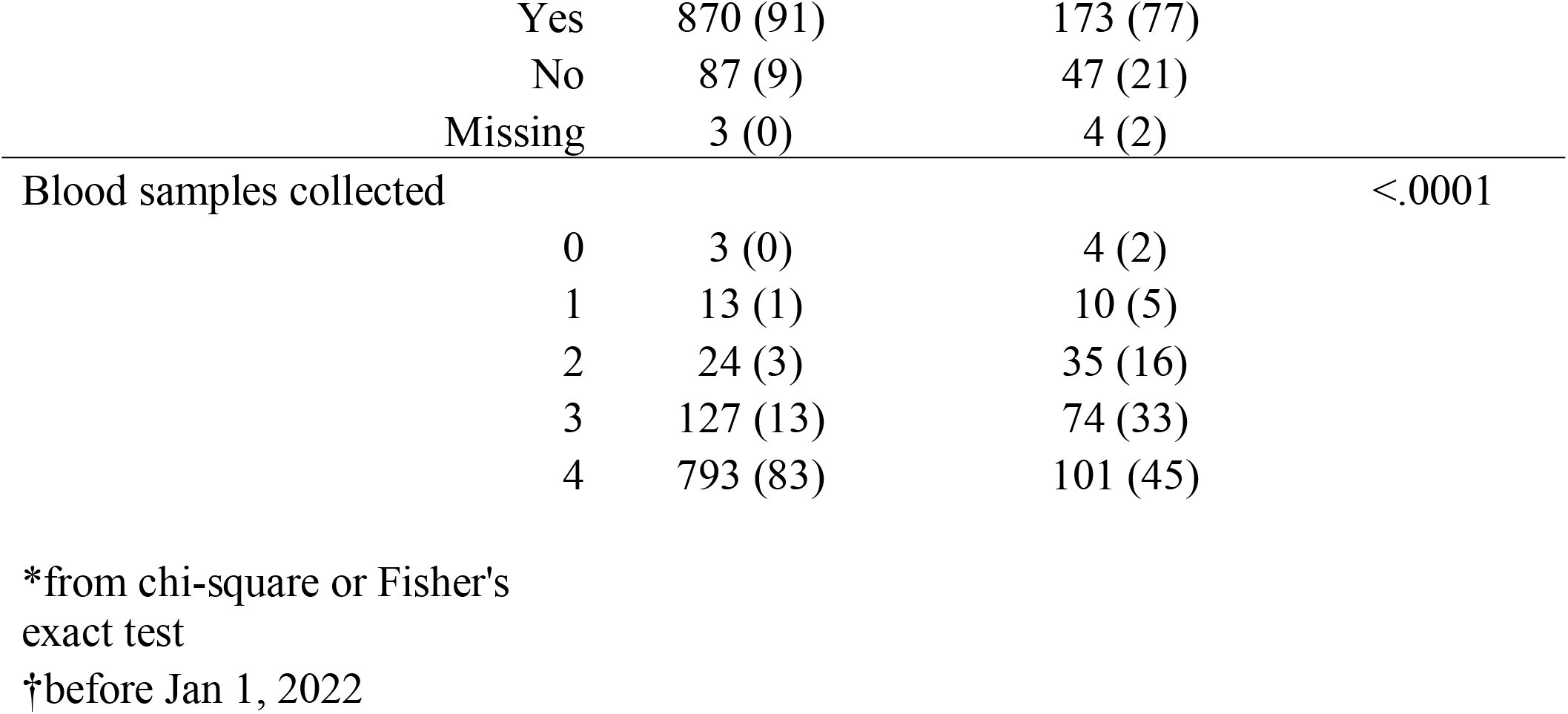
Demographics of participants eligible for SARS-CoV-2 intensive monitoring in Managua Nicaragua, March 2020-June 2022.

Over half of household activations (n=164, 55%) occurred from March 2021-December 2021, a period when multiple variants circulated, and delta predominated.[24] Additionally, there were 29 (10%) participating households in March 2020-Febuary 2021 and 104 (35%) households in January 2022-June 2022. Overall, transmission occurred in 72.4% of households (Figure 1). There were a greater proportion of primary cases that were 20-64 years old in households that had transmission compared to those where no transmission occurred (52% vs 37%) although the overall age group distribution was not significantly different (p-value: 0.0531). There were no differences in sex, bedroom- and bed-sharing, number of prior SARS-CoV-2 infections, SARS-CoV-2 vaccinations, or symptoms between primary cases of households with and without transmission (Supplemental Table 1, 2).

**Figure 1.**
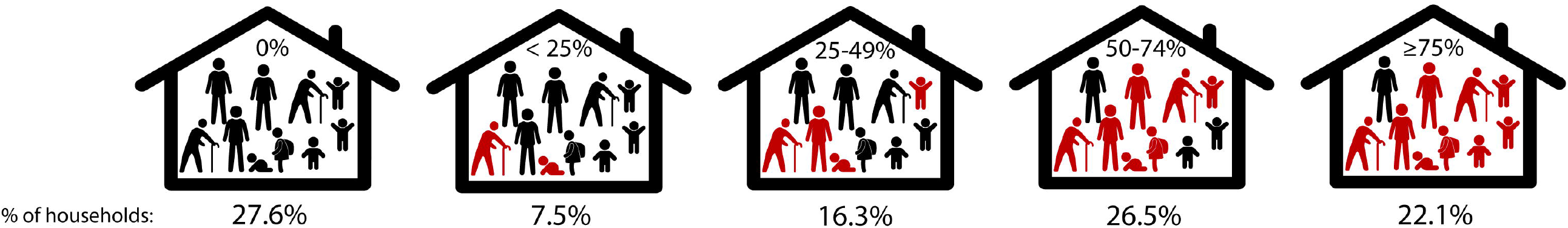
Proportion of activated households with SARS-CoV-2 transmission.

The overall estimated household SAR was 13.0% (95% CI: 11.7%, 14.6%). The estimated household SAR was smaller for larger households (8.3% compared to 15.4% for households with 10+ and 2-5 members respectively). Children (ages 5-10), and adults and adolescents (ages 11+) were much more likely to infect others compared to young children (ages 0-4) (RR of 4.20 (95% CI 1.55, 11.35) and 6.64 (95% CI: 2.59, 16.99) respectively). In absolute terms, the difference in the secondary attack rates between young children, and adults and adolescents was 11.9% (SAR of 3.2% vs 15.1%). However, there was no difference in the risk of being infected by age. Symptomatic infectious individuals were 24.37 times (95% CI: 7.80, 76.14) more likely to transmit the virus compared to asymptomatic individuals, with an absolute difference in the probability of transmission of 15.6% (SAR of 16.7% vs 1.1%). Prior SARS-CoV-2 infection was associated with protection against infection (RR=0.53, 95% CI: 0.39, 0.72) (Figure 2).

**Figure 2.**
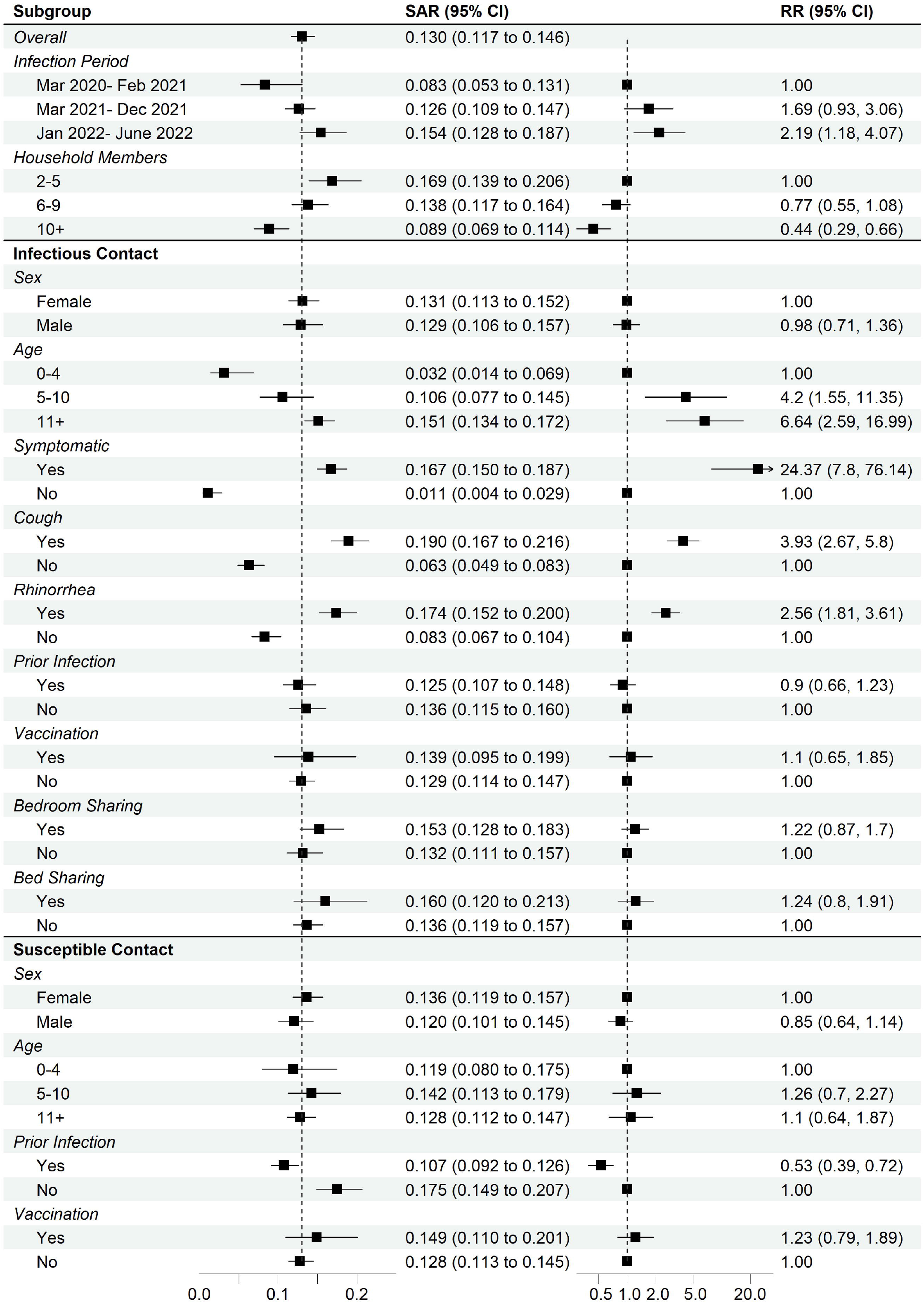
Estimated secondary attack risk and rate ratios.

For infected young children, we observed no difference by symptom status in the risk of transmitting the virus. For both infectious children and adults and adolescents, the probability of transmission was lower for asymptomatic compared to symptomatic presentation (14.5% vs 1.1% and 19.3% vs 0.4%). We note that prior infection was not associated with the probability of transmission in all age groups. Prior infection was only associated with protection against infection for adults and adolescents (Figure 3). Thus, while individuals that were previously infected were less likely to be reinfected, when reinfected they were just as likely to transmit.

**Figure 3.**
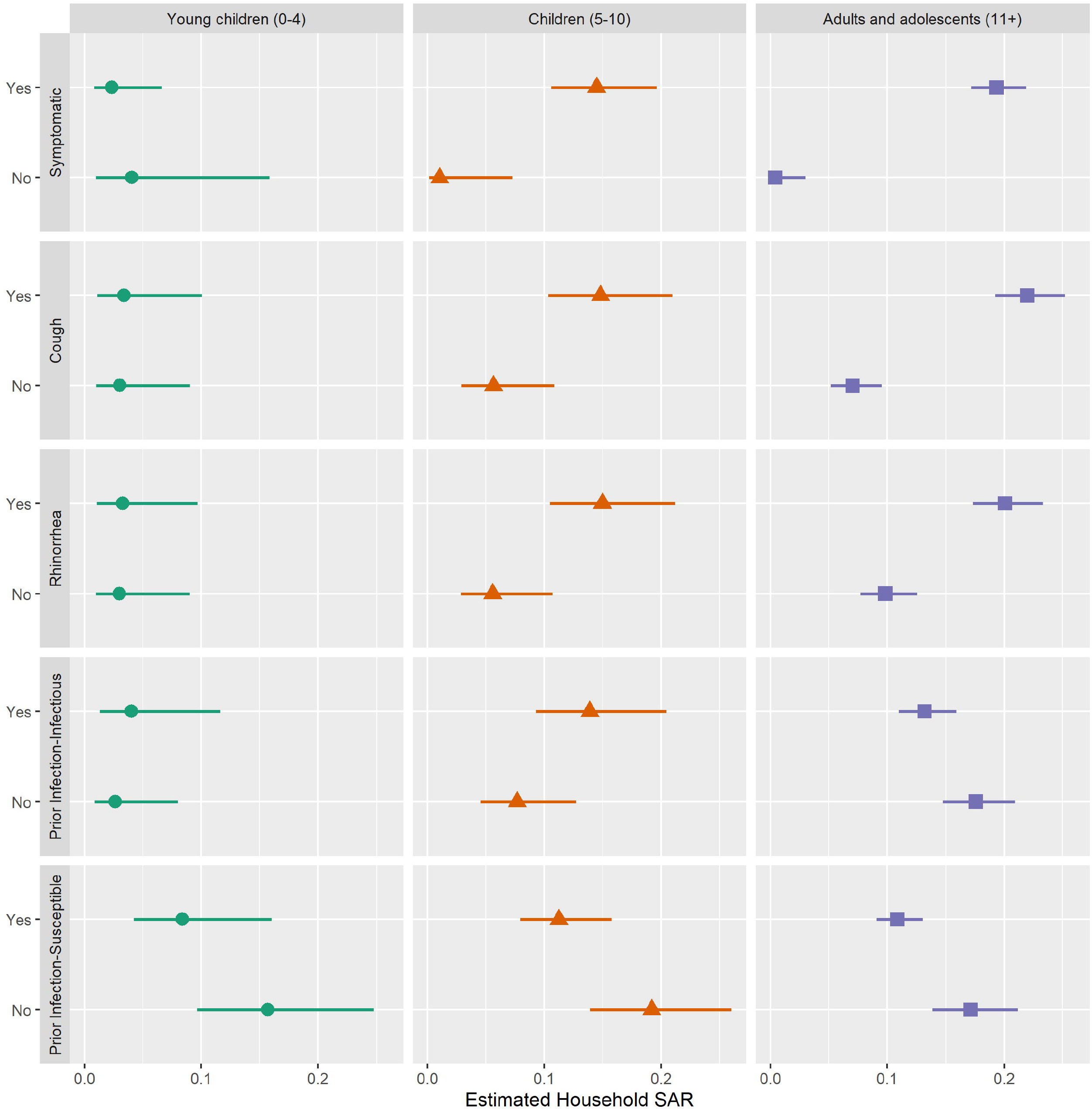
Secondary attack risk stratified by age.

Consistent with the pre-Omicron era results, during the omicron variant era, the risk of transmission was higher for symptomatic individuals (RR= 14.77, 95% CI: 3.12, 70.03) and did not vary by vaccination, and bed- or bedroom-sharing; additionally, risk of infection did not vary by age. Prior infection was still associated with protection against infection (RR=0.25, 95% CI: 0.11, 0.56). However, the risk of transmission did not vary by age as it did in the overall results. The risk of transmission was lower for males compared to females (RR= 0.30, 95% CI: 0.15,0.61) (Supplemental Figure 3, 4).

To examine the effect of our assumptions on our estimates, we varied the incubation, latency, and infectious parameters (Supplemental Figure 5). Overall, there were minor differences in the estimated SARs; however, our main findings held. To examine the effect of non-participation, we reran models limiting to households where all members participated. The overall SAR was slightly higher, but there were no differences in the direction of the association age, infection-induced immunity, or any other variable (Supplemental Figure 6).

## Discussion

We estimated the household SARS-CoV-2 SAR for a large community-based prospective cohort study in Managua, Nicaragua; to our knowledge, this is the first study that compares the association between infection-induced immunity and household SAR. We observed a decreased risk of infection for adults and adolescents who had a prior SARS-CoV-2 infection, but this was not observed among children. While estimated household SARs were much lower when the infectious contact was asymptomatic, this was not observed among young children. These results suggest distinct immune responses to natural SARS-CoV-2 infection between younger and older participants that may impact transmission dynamics.[25, 26]

Although we expected infection-induced immunity to be associated with a lower probability of transmission because of the association with decreased shedding duration and viral load,[4] this did not occur. When infected, individuals with and without infection-induced immunity had the same probability of transmission. However, these results are not inconsistent; decreased shedding duration may have little impact on household transmission of SARS-CoV-2 where household members have repeated close contact with each other early in illness. Outside of the household, decreased shedding and viral load likely leads to decrease in transmission as contact with others is likely shorter and less frequent.

During the period of the spread of the omicron variant, the results were similar to the overall findings, albeit with generally higher probability of transmission. Infection-induced immunity was still associated with protection against infection. Surprisingly, risk of transmission did not vary by age. These differences may suggest changing SARS-CoV-2 dynamics due to the omicron variant. [15, 27]

While a reduction in SARS-CoV-2 transmission for pre or asymptomatic compared to symptomatic infectious individuals has been previously noted [14, 28] and SARS-CoV-2 transmission from children compared to adults is less common [28, 29], we show that the presence of symptoms in young children is not associated with infectiousness. Thus, the increased likelihood of asymptomatic presentation of children infected with SARS-CoV-2 does not account for the differences in infectiousness between adults and children.[29]

The overall estimated household SAR of 13.0% is comparable with estimates from studies in China that also used a statistical transmission model with similar parameters (10.4% and 12.4% for incubation period of 5 days and a 13-day infectious period).[14, 30] However, studies that used estimates from primarily binomial models before and after the emergence of the omicron variant estimated a higher household SAR across settings; [2, 28, 31] while many factors may explain this difference, the use of binomial models rather than statistical transmission models likely bias the estimated SAR upwards. A prior study showed that these biased estimates cannot be interpreted as the probability of transmission, and instead statistical transmission models should be used.[18]

Our study has several strengths and limitations. Strengths include close monitoring of participants inside of an ongoing cohort, which allows us to know infection histories prior to SARS-CoV-2 entering the household as well as detect mild and asymptomatic infections. Our study is also large and spans both pre-variant and variant eras. One limitation of our study is that although PCR testing occurred frequently during monitoring, it is possible that SARS-CoV-2 infections were missed and thus we may underestimate the household SAR. Second, not all household members participated in intensive monitoring and those that declined or were not available for intensive monitoring were different from those that did participate; although the proportion with detectable SARS-CoV-2 antibodies was lower among those did not participate in activation, they on average had fewer blood samples collected. The exclusion of these participants likely leads to an underestimation of the household SAR; however, when analyzing only households where all participants consented to intensive monitoring, the probability of transmission was only slightly larger. Statistical power was also limited in our analysis of the period of omicron spread. We also note that these results are from a community where most were infected with SARS-CoV-2 prior to the availability of SARS-CoV-2 vaccines. [24] Although adults in other settings may have been vaccinated before their first SARS-CoV-2 infection, most children have not been. [6-9] However, both infection then vaccination and vaccination then infection produces broad, hybrid immunity to SARS-CoV-2 with no observed differences by sequence. [32-35]

Our study highlights the differences in SARS-CoV-2 transmission between children and adolescents and adults which may impact transmission dynamics and the transition to endemicity. Infection-induced immunity is associated with protection against infection, even in the omicron variant era, but previously infected individuals were just as likely to transmit as those that had not been previously infection. At the beginning of the SARS-CoV-2 pandemic, it was established that the contribution of children to SARS-CoV-2 transmission was minor [13]. The absence of protection against infection from infection-induced immunity among children and the changing transmission dynamics from emerging SARS-CoV-2 variants suggests that children may already have more meaningful contributions to SARS-CoV-2 transmission; this contribution may further increase as new children are born without immunity to SARS-CoV-2 and increasingly represent the greatest proportion of primary cases. [1]

## Data Availability

Data may be shared with outside investigators following UM IRB approval and once appropriate institutional approvals are obtained.

## Contributions

AMF and AG contributed to the conceptualization of the manuscript. GK, RL, SO, NS, SS, MP, CB, and AB contributed to the investigation for the manuscript. AMF conducted the statistical analysis in consultation with EK. AS was responsible for data curation. AMF and AG wrote the original draft of the manuscript and all co-authors contributed to the review and editing of the manuscript. AF, AS, GK, AB, and AG had access to data and verify its authenticity.

