## Supplemental Methods for "Infection-induced immunity is associated with protection against SARS-CoV-2 infection, but not decreased infectivity during household transmission"

**Supplementary methods**

Bedroom- and bed-sharing information was available from household enrollment surveys. Pairwise survival models are explicit statistical models of disease transmission that overcome weaknesses of binomial models in estimating the household SAR. Pairwise survival models can use the entire household observation period to estimate the SAR, not just infectious period of the index case, even when who-infects-who is not observed. For the pairwise regression models, each infected participant was paired with each susceptible participant during their infectious period. Participants’ infection date was considered the earliest of 1.) the date of first PCR+ or 2.) the date of symptom onset. The external infection model for the pairwise regression included all HICS participants, not just those in activated households. For both the external and internal models, we used a log-logistic contact interval distribution which generally produced better model fit (lower AIC) than the exponential or Weibull distributions.
