## Supplemental Table 1 for "Infection-induced immunity is associated with protection against SARS-CoV-2 infection, but not decreased infectivity during household transmission"

**Supplemental Table 1. Primary case characteristics by presence of household transmission**

|  | ≥ 1 Household Contact PCR+ | |  |
| --- | --- | --- | --- |
|  | No (n=84) | Yes (n=213) | p-value* |
| Pandemic Period (%) |  |  | 0.0514 |
| Mar 2020 - Feb 2021 | 14 (17) | 15 (7) |  |
| Mar 2021 - Dec 2021 | 43 (51) | 121 (57) |  |
| Jan 2022 - May 2022 | 27 (32) | 77 (36) |  |
| Female (%) | 58 (69) | 136 (64) | 0.3966 |
| Age Group (%) |  |  | 0.0531 |
| 0-4 | 12 (14) | 12 (6) |  |
| 5-10 | 16 (19) | 41 (19) |  |
| 11-19 | 21 (25) | 43 (20) |  |
| 20-64 | 31 (37) | 110 (52) |  |
| 65+ | 4 (5) | 7 (3) |  |
| Share bedroom | 62 (84) | 177 (85) | 0.7874 |
| Share bed | 43 (58) | 131 (63) | 0.459 |
| Prior SARS-CoV-2 Infections (%) |  |  | 0.5238 |
| 0 | 42 (50) | 109 (51) |  |
| 1 | 40 (48) | 93 (44) |  |
| 2 | 2 (2) | 11 (5) |  |
| Completed SARS-CoV-2 Vaccinations (%) | 10 (12) | 25 (12) | 0.9678 |
| Symptoms |  |  |  |
| Cough | 67 (80) | 168 (79) | 0.8652 |
| Rhinorrhea | 60 (71) | 162 (76) | 0.4084 |
| *from chi-square or Fisher's exact test, uncorrected |  |  |  |
