## Supplemental Table 2 for "Infection-induced immunity is associated with protection against SARS-CoV-2 infection, but not decreased infectivity during household transmission"

**Supplemental Table 2. Household contact characteristics by SARS-CoV-2 infection**

|  | PCR- Contacts (n=695) | PCR+ Contacts (n=494) | p-value* |
| --- | --- | --- | --- |
| Female | 428 (62) | 299 (61) | 0.7127 |
| Age Group (%) |  |  | 0.8073 |
| 0-4 | 63 (9) | 38 (8) |  |
| 5-10 | 151 (22) | 108 (22) |  |
| 11-19 | 157 (23) | 122 (25) |  |
| 20-64 | 282 (42) | 200 (40) |  |
| 65+ | 32 (5) | 26 (5) |  |
| Share bedroom | 595 (86) | 427 (87) | 0.6239 |
| Share bed | 438 (63) | 301 (61) | 0.4639 |
| Prior SARS-CoV-2 Infections (%) |  |  | 0.0916 |
| 0 | 245 (37) | 202 (42) |  |
| 1 | 394 (59) | 253 (54) |  |
| 2 | 31 (5) | 26 (5) |  |
| 3 | 0 | 1 (0) |  |
| Completed SARS-CoV-2 Vaccination (%) | 69 (10) | 74 (15) | 0.0083 |
| *from chi-square or Fisher's exact test, uncorrected |  |  |  |
