## Supplementary figures and images for "Infection-induced immunity is associated with protection against SARS-CoV-2 infection, but not decreased infectivity during household transmission"

### Supplemental Figure 1

**Supplemental Figure 1. Flowchart of participants by year**

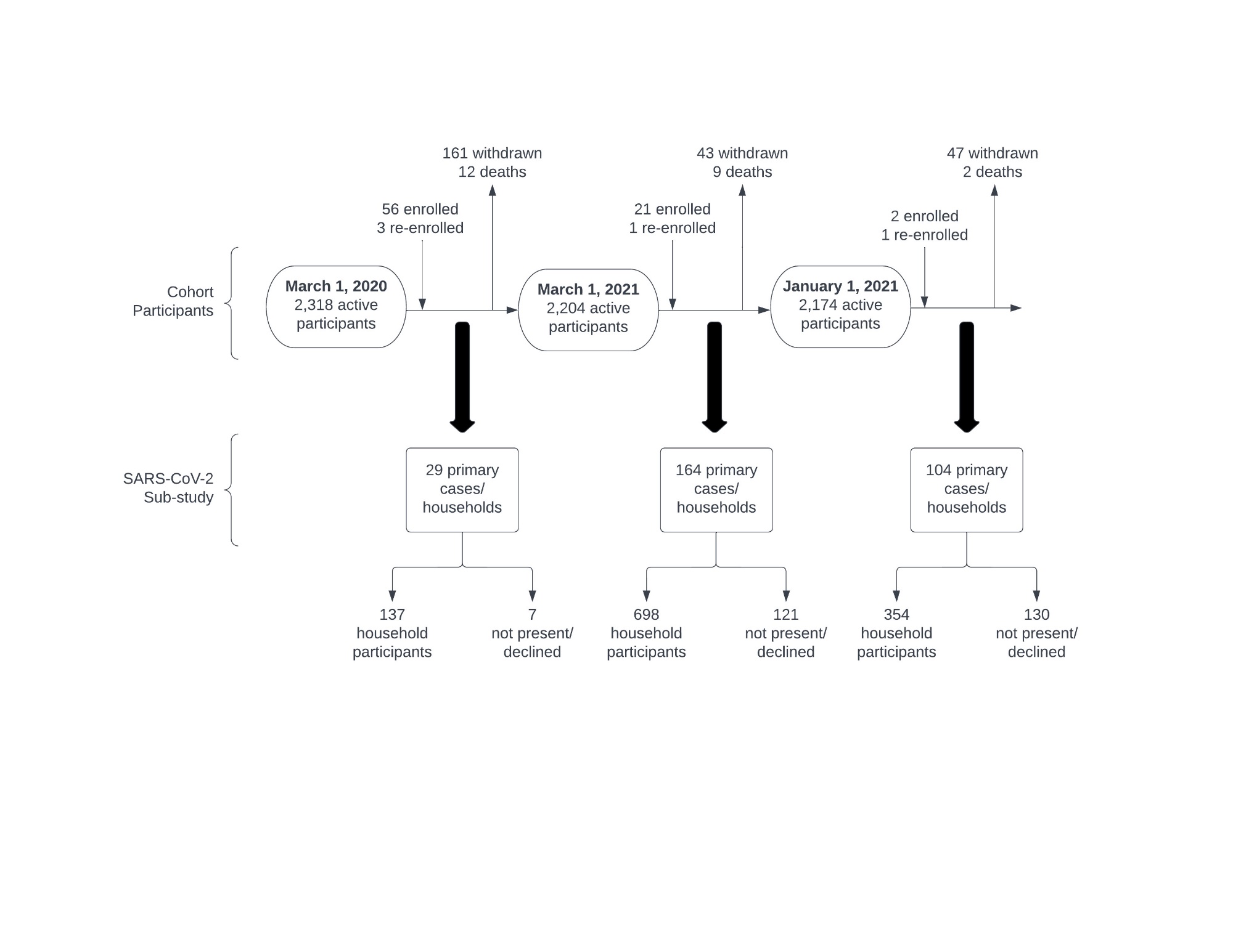

### Supplemental Figure 2

**Supplemental Figure 2. Activated households and participants**

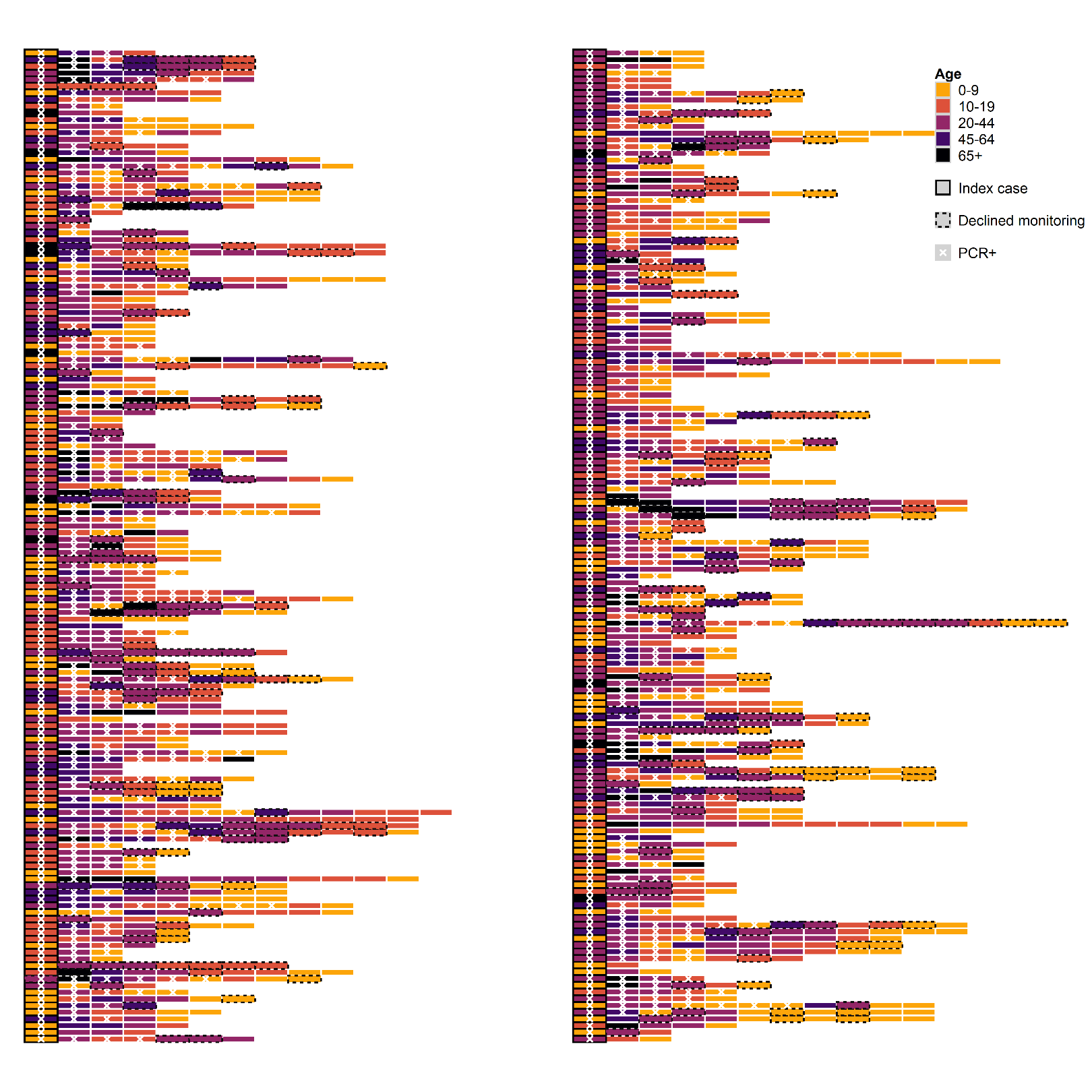

### Supplemental Figure 3

**Supplemental Figure 3. Omicron era, estimated secondary attack risk and risk ratios**

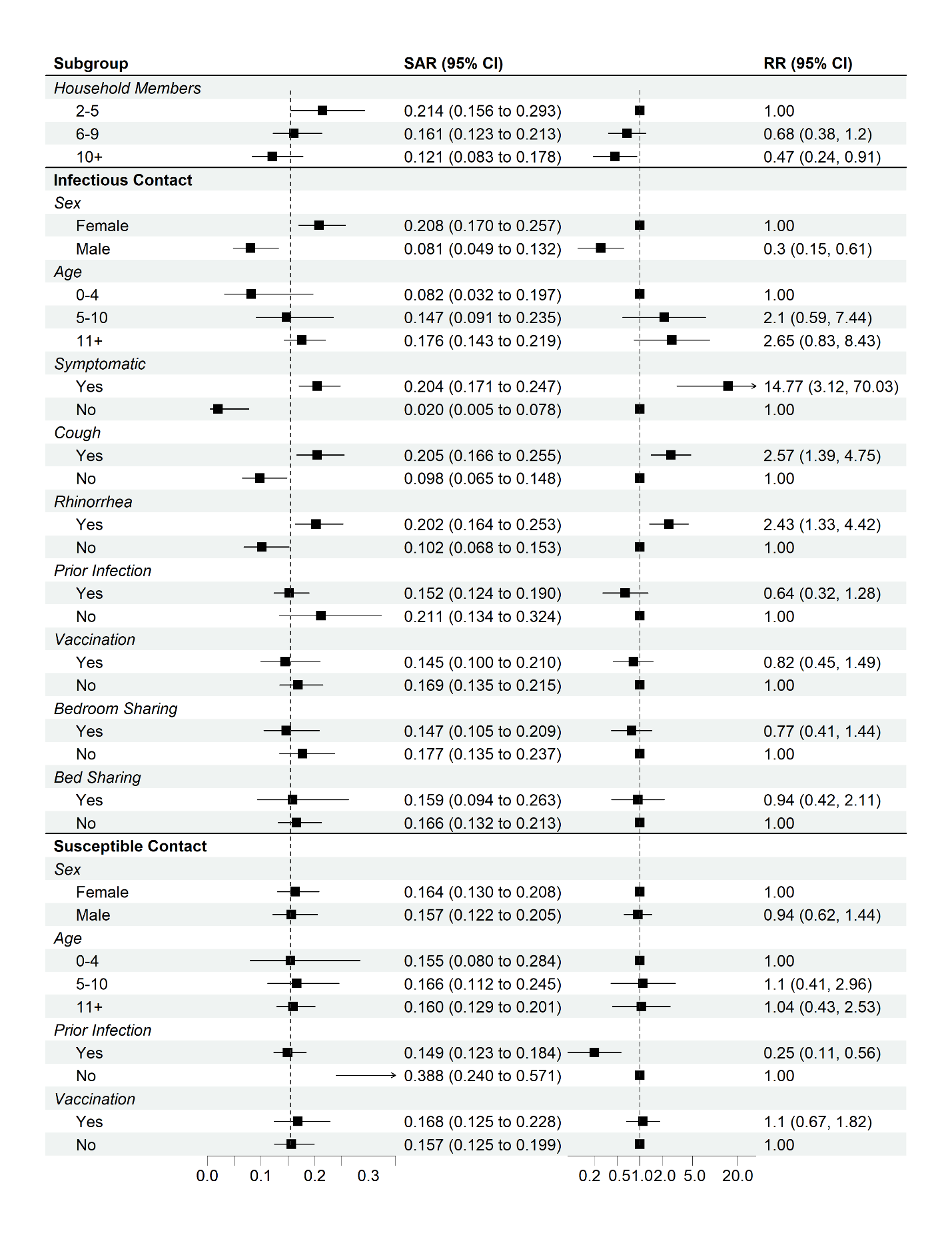

### Supplemental Figure 4

**Supplemental Figure 4. Pre-omicron era, estimated secondary attack risk and risk ratios**

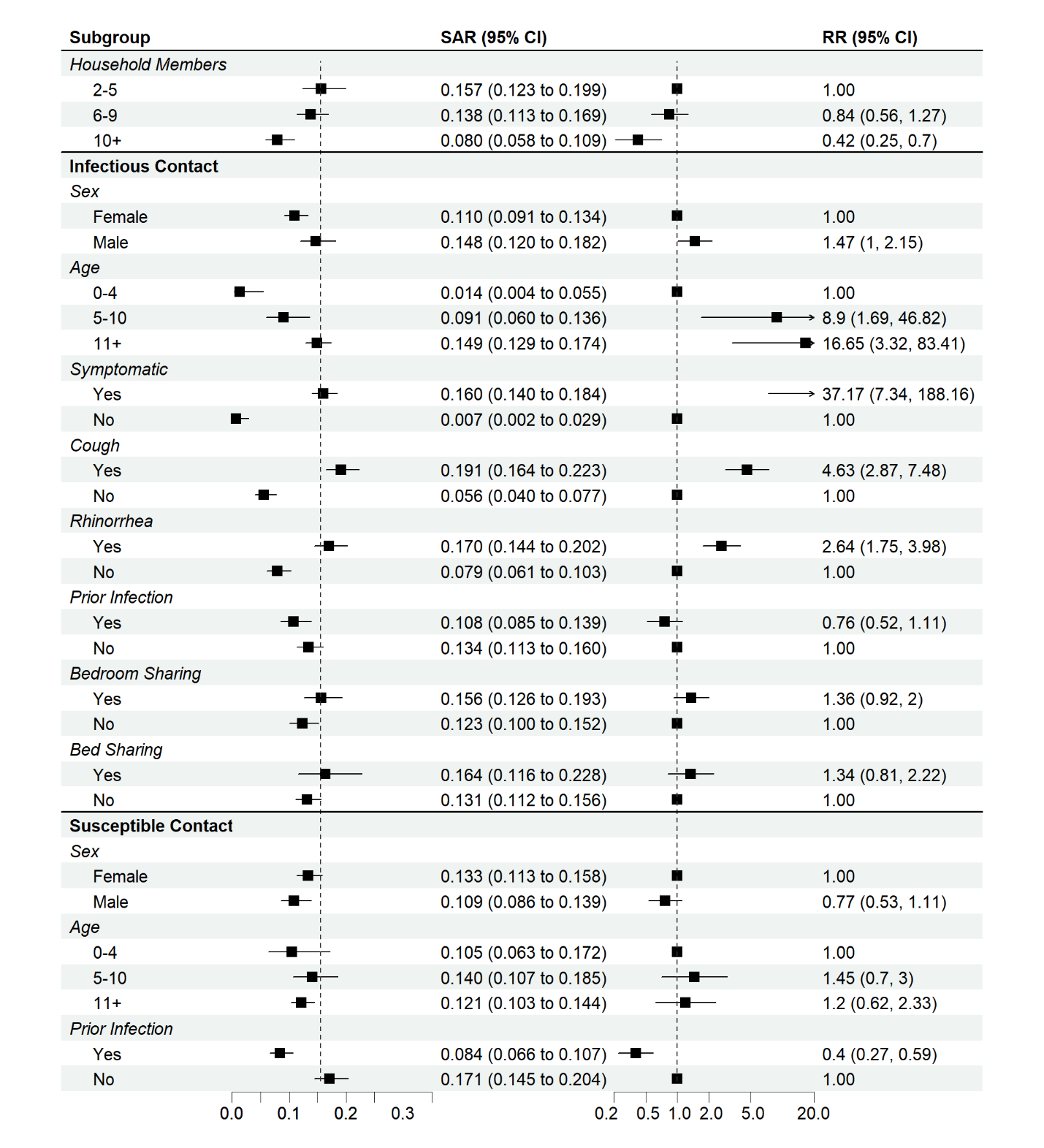
