## Supplemental Figure 5 for "Infection-induced immunity is associated with protection against SARS-CoV-2 infection, but not decreased infectivity during household transmission"

**Supplemental Figure 5. Secondary attack risk by changes in the latency, incubation, and infectivity periods**

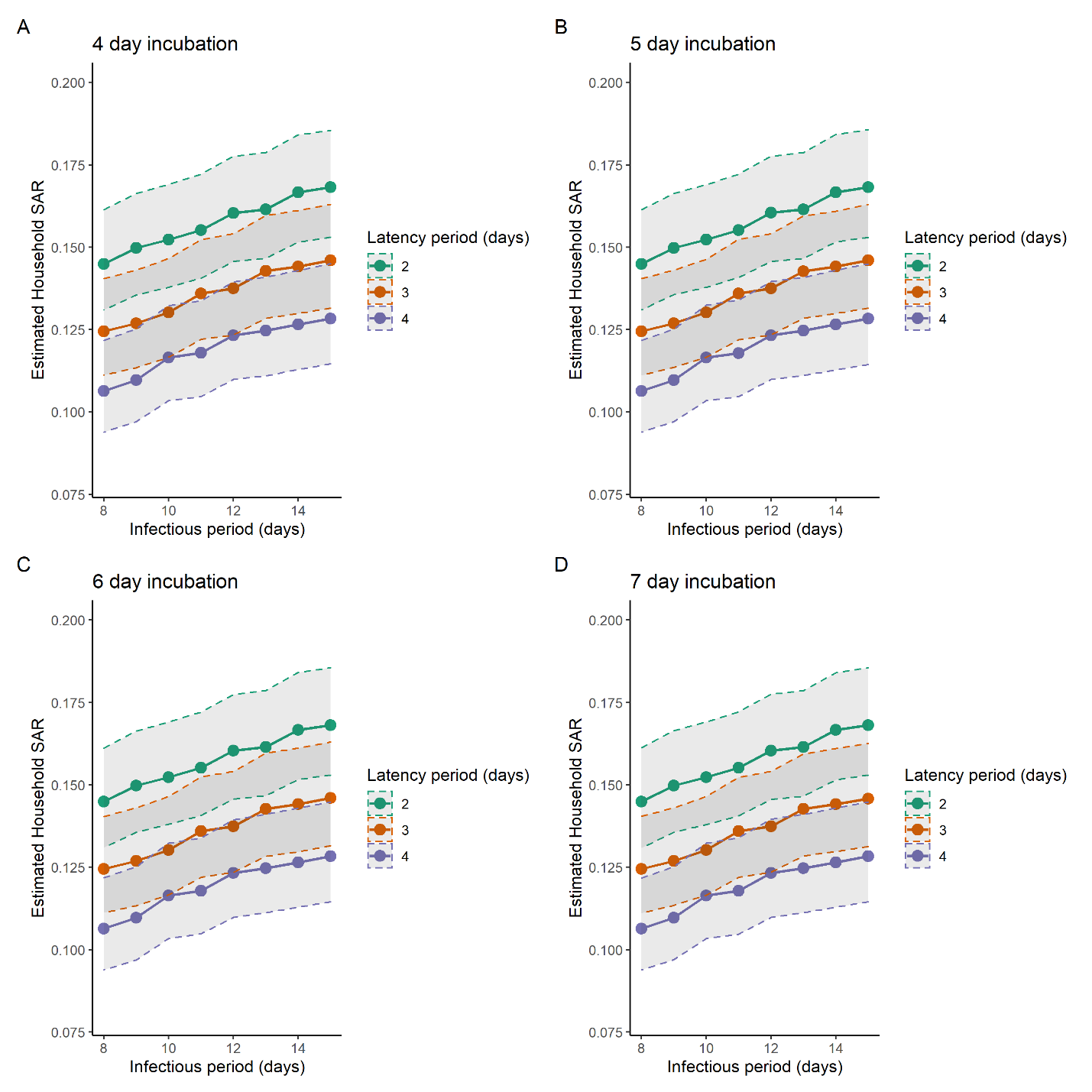
