## Supplemental Figure 6 for "Infection-induced immunity is associated with protection against SARS-CoV-2 infection, but not decreased infectivity during household transmission"

**Supplemental Figure 6. Households with complete participation, estimated secondary attack risk and risk ratios**

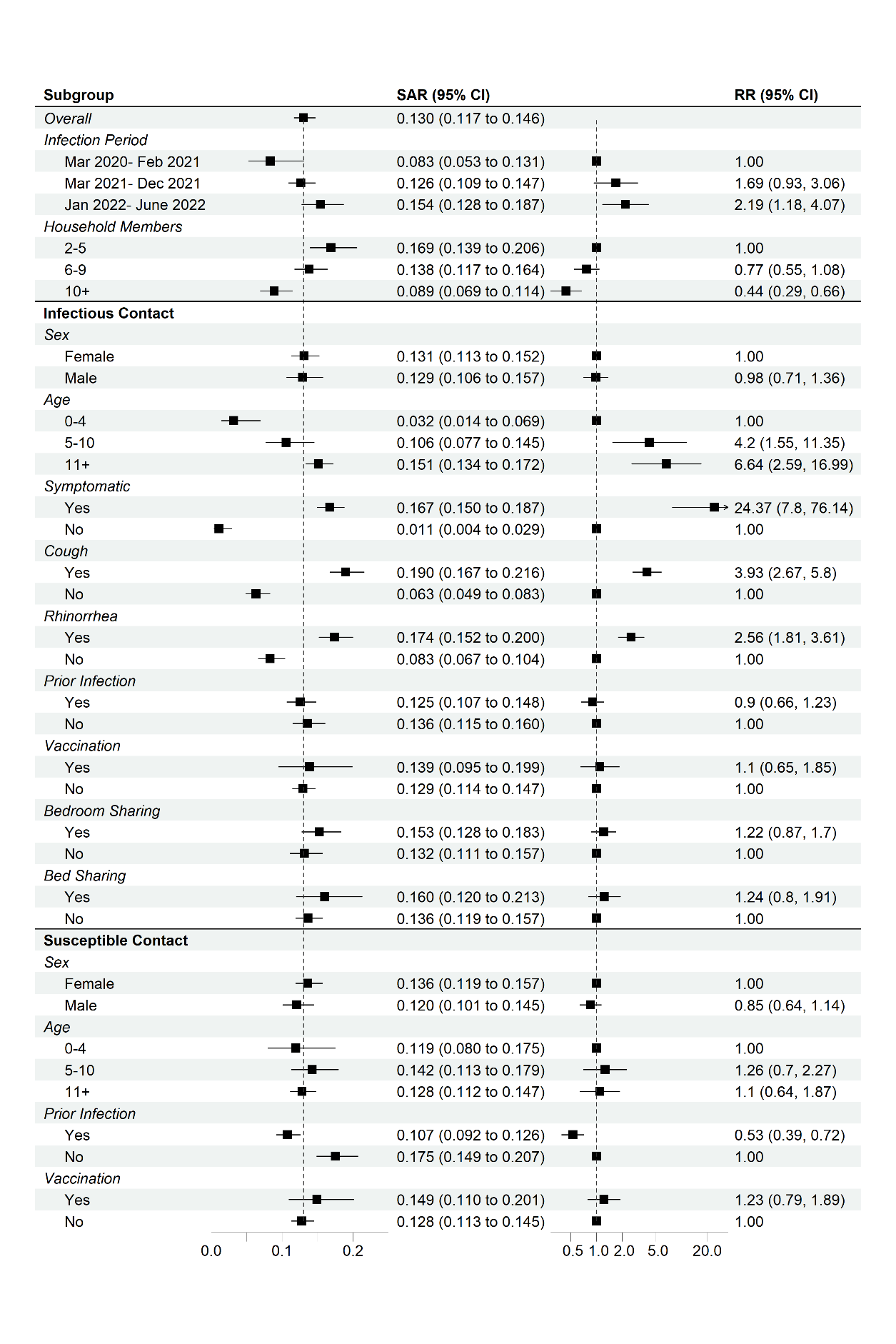
